# Assessing the clinical severity of the Omicron variant in the Western Cape Province, South Africa, using the diagnostic PCR proxy marker of RdRp target delay to distinguish between Omicron and Delta infections – a survival analysis

**DOI:** 10.1101/2022.01.13.22269211

**Authors:** Hannah Hussey, Mary-Ann Davies, Alexa Heekes, Carolyn Williamson, Ziyaad Valley-Omar, Diana Hardie, Stephen Korsman, Deelan Doolabh, Wolfgang Preiser, Tongai Maponga, Arash Iranzadeh, Sean Wasserman, Linda Boloko, Greg Symons, Peter Raubenheimer, Abraham Viljoen, Arifa Parker, Neshaad Schrueder, Wesley Solomon, Petro Rousseau, Nicole Wolter, Waasila Jassat, Cheryl Cohen, Richard Lessells, Robert J Wilkinson, Andrew Boulle, Nei-yuan Hsiao

## Abstract

**Background:** Emerging data suggest that SARS-CoV-2 Omicron variant of concern (VOC)is associated with reduced risk of severe disease. The extent to which this reflects a difference in the inherent virulence of Omicron, or just higher levels of population immunity, is currently not clear.

**Methods:** RdRp target delay (RTD: a difference in cycle threshold value of RdRp - E > 3.5) in the Seegene Allplex™ 2019-nCoV PCR assay is a proxy marker for the Delta VOC. The absence of this proxy marker in the period of transition to Omicron was used to identify suspected Omicron VOC infections.

Cox regression was performed for the outcome of hospital admission in those who tested positive for SARS-CoV-2 on the Seegene Allplex™ assay from 1 November to 14 December 2021 in the Western Cape Province, South Africa, public sector. Vaccination status at time of diagnosis, as well as prior diagnosed infection and comorbidities, were adjusted for.

**Results:** 150 cases with RTD (proxy for Delta) and 1486 cases without RTD (proxy for Omicron) were included. Cases without RTD had a lower hazard of admission (adjusted Hazard Ratio [aHR] of 0.56, 95% confidence interval [CI] 0.34-0.91). Complete vaccination was protective of admission with an aHR of 0.45 (95%CI 0.26-0.77).

**Conclusion:** Omicron has resulted in a lower risk of hospital admission, compared to contemporaneous Delta infection in the Western Cape Province, when using the proxy marker of RTD. Under-ascertainment of reinfections with an immune escape variant like Omicron remains a challenge to accurately assessing variant virulence.

## Background

With the rapid global spread of the Omicron (B.1.1.529) SARS-CoV-2 variant of concern (VOC), understanding the clinical implications of this new VOC is critical [1]. Emerging data from both South Africa and the United Kingdom suggest that it is associated with a reduced risk of severe disease [2–7]. The extent to which this reflects a difference in the inherent virulence of Omicron, or just higher levels of population immunity, due to previous infection and/or vaccination, is currently not clear.

In November 2021 in the Western Cape Province, South Africa, following a period of very low incidence, Omicron rapidly replaced Delta (B.1.617.2) as the dominant variant and drove the fourth wave of infections. Omicron is known to result in S-gene target failure (SGTF) on the Thermo Fisher Taqpath™ PCR assay [8]. Unfortunately, too little routine diagnostic testing is done using this assay in the Western Cape National Health Laboratory Service (NHLS) to power an SGTF analysis.

RdRp target delay (RTD) is a proxy marker for the Delta variant on routine diagnostic testing with the Seegene Allplex™ 2019-nCoV PCR assay, similar in concept to S-gene target failure. RTD has a 93.6% sensitivity and 89.7% specificity in detecting the Delta variant when compared to genomic sequencing [9]. This proxy marker has previously successfully been used to assess the association of the Delta variant and mortality in the third wave [10].

As the Seegene Allplex™ assay is widely used by the Western Cape NHLS, and as Omicron rapidly displaced the Delta variant with minimal other variants detected, the absence of RTD in Seegene Allplex™ cases can be used to identify likely Omicron infections during the replacement period. The absence of RTD tracks well with SGTF in the province (Figure 1), as well as genomic sequenced data [1,11]. We used this proxy marker to assess the clinical severity associated with Omicron infection.

**Figure 1:**
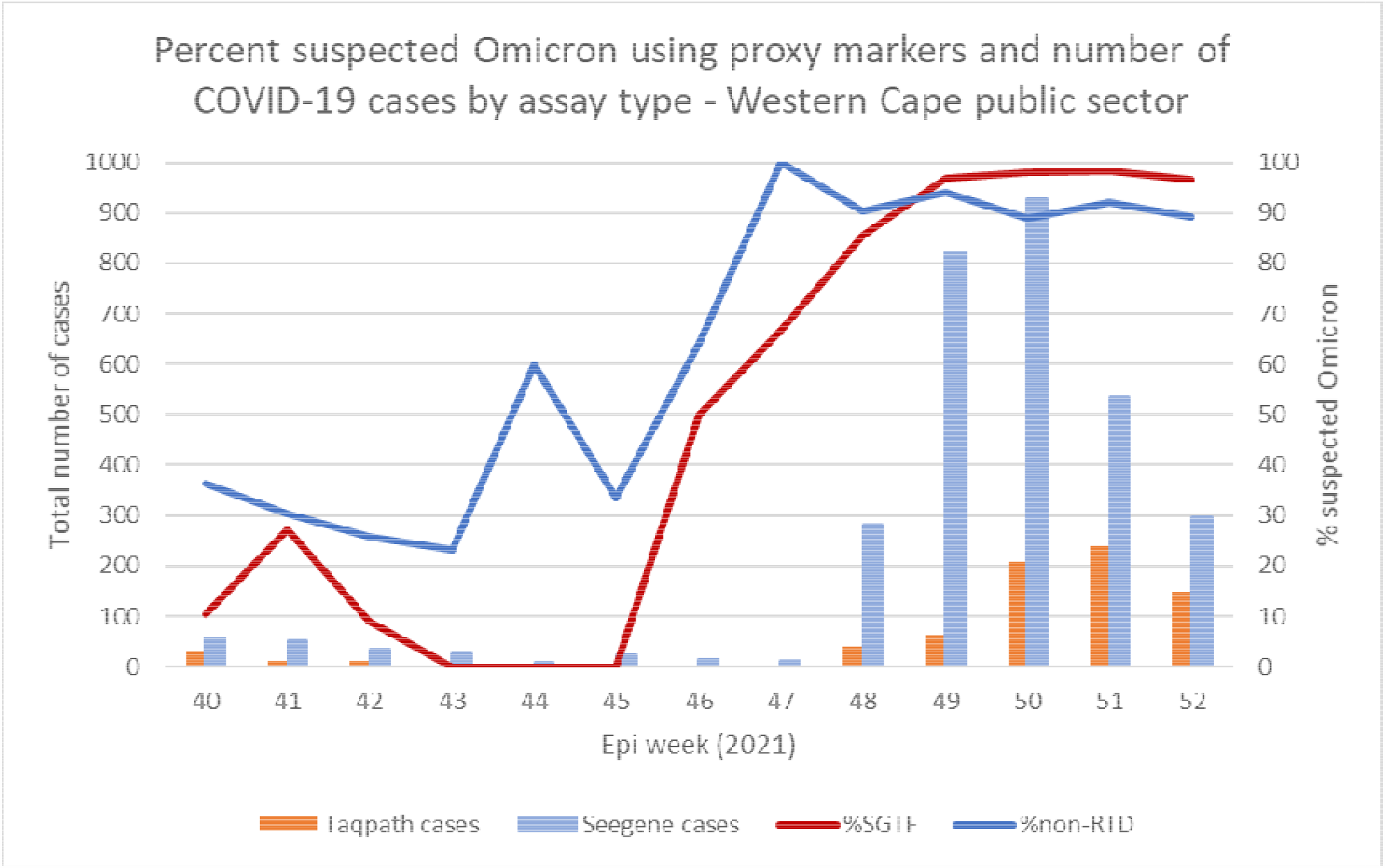
Percent suspected Omicron using the proxy markers of S-gene target failure in Thermo Fisher Taqpath™ cases and non-RdRp target delay in Seegene Allplex™ assay cases, in the Western Cape Province public sector adults.

## Methods

RTD is defined as a difference in cycle threshold value of RdRp gene target - E gene target > 3.5 in the Seegene Allplex™ 2019-nCoV PCR assay [9]. Cases diagnosed using this assay have previously been found to be representative of cases diagnosed by any PCR assay in the Western Cape Province public sector [10].

We included all COVID-19 cases, aged 15 years and older, diagnosed on the Seegene Allplex™ 2019-nCoV diagnostic PCR assay in the Western Cape Province public sector from 1 November to 14 December 2021, a period when both Delta and Omicron were co-circulating and other lineages were negligible [1,11]. Follow-up ended 6 January 2022. Children were excluded from this analysis as their testing and admission patterns are different compared to adults.

The Western Cape Provincial Health Data Centre (PHDC) collates all available electronic health data on public sector patients, including COVID-19 test results, comorbidities and hospital admissions. If an individual tested positive for SARS-CoV-2 14 days before admission, or 7 days after the date of admission, and was not admitted to a specialized psychiatric and rehabilitation facilities, it was defined as COVID-19 related hospital admission.

We undertook a survival analysis using Cox regression, assessing time from date of diagnosis to date of hospital admission, which was our outcome of interest. Those whose date of admission was on or before the date of diagnosis were assigned one day of follow-up. Intensive Care Unit (ICU) admission or mortality were not assessed as too few of these severe outcomes had occurred. We adjusted for age, sex, known comorbidities, prior diagnosed infection (two positive COVID-19 tests more than 90 days apart) and vaccination status at time of diagnosis. Vaccination status was determined by linking COVID-19 cases with the national Electronic Vaccination Data System through national identifiers in the PHDC. Complete vaccination was defined ≥28 days post-vaccination with Janssen/Johnson & Johnson (Ad26.COV2.S), or ≥14 days post second dose of Pfizer–BioNTech (BNT162b2). Patients were deemed partially vaccinated from the day after their (first) vaccine dose until meeting criteria for complete vaccination. Analyses were conducted using Stata 13.1.

The study was approved by the University of Cape Town Research Ethics Committee (HREC 460/2020).

## Results

We included 150 cases with RTD (proxy for Delta) and 1486 cases without RTD (proxy for Omicron/non-Delta). Patient demographic characteristics and comorbidities were similar in both groups (Table 1). The median age in both groups was 33 years (IQR 25-44 in those with RTD; IQR 26-44 in those without). The proportion of cases that were fully vaccinated in both groups was also similar, although more infections following partial vaccination were seen in those without RTD.

**Table 1:** Characteristics of included cases, according to presence or absence of RdRp target delay, as well as adjusted hazard ratios (aHR) for the outcome of admission in adults in the Western Cape Province public sector, 1 November to 14 December 2021.

|  |  | RTD present | RTD absent | Cox regression for outcome of admission |  |
| --- | --- | --- | --- | --- | --- |
|  |  | 150 | 1486 | 1636 (122 admissions) |  |
|  |  | n (%) | n (%) | adjusted HR | 95% CI |
| RTD | Present ("Delta") |  |  | Ref |  |
|  | Absent ("Omicron") |  |  | 0.56 | 0.34; 0.91 |
| Sex | Female | 80 (53.3%) | 860 (57.9%) | Ref |  |
|  | Male | 70 (46.7%) | 626 (42.1%) | 1.03 | 0.71; 1.48 |
| Age category | 15-29 years | 57 (38%) | 559 (37.6%) | Ref |  |
|  | 30-39 years | 43 (28.7%) | 447 (30.1%) | 0.92 | 0.56; 1.50 |
|  | 40-49 years | 21 (14%) | 216 (14.5%) | 1.12 | 0.62; 2.02 |
|  | 50-59 years | 15 (10%) | 166 (11.2%) | 0.91 | 0.46; 1.80 |
|  | 60-69 years | 6 (4%) | 60 (4%) | 1.60 | 0.73; 3.49 |
|  | ≥70 years | 8 (5.3%) | 38 (2.6%) | 1.27 | 0.56; 2.89 |
| Comorbidity§ | HIV positive | 21 (14%) | 142 (9.6%) | 1.29 | 0.72; 2.30 |
|  | Diabetes | 7 (4.7%) | 77 (5.2%) | 1.30 | 0.68; 2.46 |
|  | Tuberculosis (current) | 4 (2.7%) | 17 (1.1%) | 5.15 | 2.44; 8.89 |
|  | Hypertension | 19 (12.7%) | 149 (10%) | 1.78 | 0.99; 3.19 |
|  | Chronic Kidney Disease | 4 (2.7%) | 23 (1.6%) | 4.46 | 2.24; 8.89 |
|  | COPD / Asthma | 6 (4%) | 79 (5.3%) | 2.26 | 1.29; 3.96 |
| Prior infection | None | 134 (89.3%) | 1313 (88.4%) | Ref |  |
|  | Yes | 16 (10.7%) | 173 (11.6%) | 0.00 |  |
| Vaccination † | None | 114 (76%) | 1081 (72.7%) | Ref |  |
|  | Partial | 3 (2%) | 93 (6.3%) | 1.22 | 0.63; 2.36 |
|  | Complete | 33 (22%) | 312 (21%) | 0.45 | 0.26; 0.77 |

§ The reference group for the aHR here is the absence of that specific comorbidity.
† Fully vaccinated was defined as ≥28 days post-vaccination with Janssen/Johnson & Johnson or ≥14 days post second dose of Pfizer–BioNTech. Patients were deemed partially vaccinated from the day after their (first) vaccine dose until meeting criteria for complete vaccination.

The proportion of cases with a documented reinfection was 11% for both those with and without RTD. By using a stricter definition of RTD (i.e., requiring a larger difference in Ct values), a similar proportion of RTD cases that had documented reinfections were found (Supplementary Table 1).

21 cases (14%) were admitted to hospital in those with RTD, while 101 (6.8%) were admitted in those without RTD. Amongst those not admitted, 189 (12.5%) had a documented previous infection, while amongst those admitted, none had a documented previous infection and so we could not calculate the extent of protection from prior diagnosed infection against admission, although there appears to be benefit. The cases without RTD (i.e. suspected Omicron cases) had a lower hazard of admission (adjusted Hazard Ratio [aHR] of 0.56, 95% confidence interval [CI] 0.34-0.91) (Table 1), after adjusting for age, sex, prior diagnosed infection, vaccination status at time of diagnosis and known comorbidities, when compared to suspected Delta cases. Complete vaccination was protective of admission with an aHR of 0.45 (95%CI 0.26-0.77).

The proportional hazards assumption was tested and found to have held for each variable and the model as a whole (see Supplementary Table 2).

## Discussion

Using the proxy marker of RTD absence, suspected Omicron cases were associated with a lower risk of hospital admission when compared to contemporaneous suspected Delta cases.

A study from the South African National Institute of Communicable Diseases found that using SGTF suspected Omicron cases had a lower odds of being admitted to hospital compared to non-SGTF infections (adjusted odds ratio [aOR] 0.2, 95%CI 0.1-0.3) [2]. However, this analysis was not able to adjust for vaccination status at time of diagnosis, potentially explaining the greater reduction in observed disease severity compared to our study. Similar contemporaneous SGTF studies from the UK have also found that SGTF was associated with a lower risk of hospitalisation, with an adjusted observed/expected ratio of 0.32 (95% CI 0.19, 0.52) [4] and a 40-45% reduction in risk of admission, which is similar to our result [5].

Several studies have also found less severe disease in the fourth Omicron-driven wave compared to the third wave caused by Delta in South Africa [3,6,7], but as they are comparing non-contemporaneous cases it is difficult to account for the effect of the vaccination programme, which started too late to provide significant protection for the third wave [12], as well as the fact that more individuals were previously infected, and thus had some immunity at the start of the fourth wave compared to the third.

Anecdotally, this fourth wave has resulted in a relatively large proportion of admissions where COVID-19 was incidentally diagnosed [3]. It is unfortunately not possible to tease out the primary indication for admission from our routine data and we were unable to assess severe outcomes due to the very small numbers of cases tested on the Seegene Allplex™ assay who were recorded as having steroids prescribed electronically, being admitted to ICU or dying. The fact that full vaccination still provided substantial protection against admission, even with the contamination of incidental admissions, suggests that vaccination provides very strong protection against admission in the face of the Omicron variant, and that some of the admissions were likely due to COVID-19 disease. Vaccination itself might also be a proxy marker for higher socio-economic status, access to health care or good health seeking behaviour, such as adherence to chronic medications, and as such might confer some protection against hospital admissions, irrespective of COVID-19.

Omicron is associated with an increased risk of reinfections [13]. Previous infection in itself is protective against severe disease [14]. In this analysis, no cases with a documented reinfection required hospital admission. But seroprevalence studies indicate only a small fraction of total COVID-19 cases in the Western Cape Province are laboratory confirmed [15], particularly when there have been testing restrictions during epidemic surges. Under-ascertainment of prior diagnosed infection is therefore likely, and this could considerably bias measures of disease severity for an immune escape variant compared to a variant that is not associated with reinfection. The extent of this residual confounding i.e., the contribution of this under-ascertainment of reinfections to the milder disease severity seen with Omicron, is thus still uncertain in our context of very high rates of unconfirmed prior infection. At the same time, if we are diagnosing a smaller proportion of Omicron cases by missing the more mild or asymptomatic infections, or if the Omicron variant results in more incidental hospital admissions and outcome misclassification, this could bias our findings in the opposite direction, and falsely elevate the disease severity seen with this variant.

Additional limitations to this study are the low numbers of SARS-CoV-2 infections during the Delta to Omicron transition period, and that we used a proxy marker for Omicron and not whole genome sequencing, as only limited sequencing is feasible in our setting. A proxy marker on routine diagnostic testing is likely to result in some variant misclassification, which is evidenced by the high rate of reinfections seen in RTD cases. Attempts to narrow the definition of RTD were not able to resolve this. In addition, this analysis like many others, only compares disease severity between Omicron and Delta. Little is known of the severity of Omicron when compared to the ancestral strain and other non-Delta VOCs.

## Conclusion

Omicron has resulted in a lower risk of hospital admission, when compared to contemporaneous Delta cases using the proxy marker of RTD in the Western Cape Province. Vaccination and documented previous infection are highly protective of hospital admission. Under-ascertainment of reinfections with an immune escape variant like Omicron remains a challenge to accurately assessing variant virulence.

## Data Availability

All data produced in the present study are available upon reasonable request to the authors.

---

**Supplementary Table 1:**
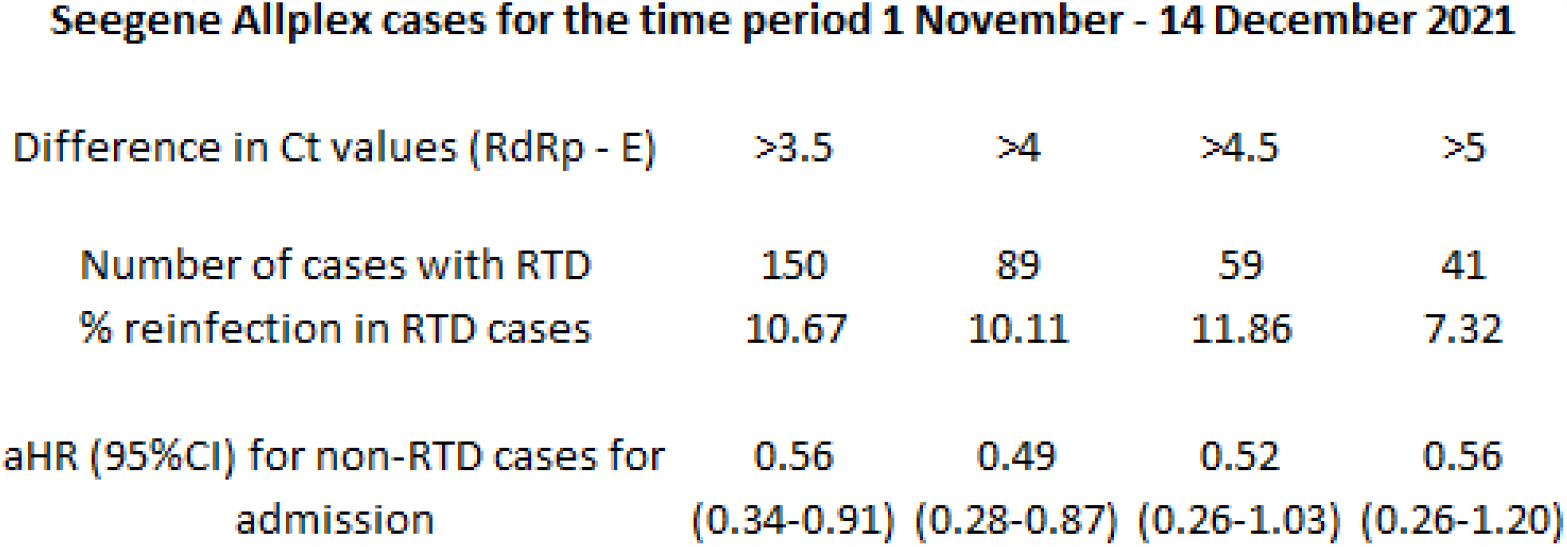
Adjusting the difference in Ct values (RdRp – E), and the resultant adjusted Hazard Ratios for suspected Omicron cases being admitted, using the same survival analysis model as the main analysis. Using the original definition of RTD (>3.5), 16 RTD cases with reinfections (10.67%) were included in the main analysis. The proportion of RTD cases that were reinfections was 2.6, 2.1 and 0.8% in the months of August, September and October 2021 respectively. By narrowing the definition of RTD to require a larger difference in Ct values, less “Delta” cases were classified as such, but the aHR for suspected Omicron cases and admission remained similar, although the confidence intervals did widen. The stricter definition of RTD, however, was not able decrease the proportion of cases with reinfection. In November, when Delta was still dominant, there were very low case numbers. After Omicron arrived, there was a massive increase in case numbers. A small proportion of misclassification of these higher numbers of Omicron cases is therefore able to markedly change the proportion of reinfection seen in the RTD cases.

**Seegene Allplex cases for the time period 1 November - 14 December 2021**
| Difference in Ct values (RdRp - E) | >3.5 | >4 | >4.5 | >5 |
| --- | --- | --- | --- | --- |
| Number of cases with RTD | 150 | 89 | 59 | 41 |
| % reinfection in RTD cases | 10.67 | 10.11 | 11.86 | 7.32 |
| aHR (95%CI) for non-RTD cases for admission | 0.56<br>(0.34-0.91) | 0.49<br>(0.28-0.87) | 0.52<br>(0.26-1.03) | 0.56<br>(0.26-1.20) |

**Supplementary Table 2:** Assessing the proportional hazards assumption on the basis of Schoenfeld residuals, for each variable and the model as whole. Although the p values for CKD and complete vaccination were low, the log-log plots (Supplementary Figure 1) showed no gross violation.

|  |  | rho | chi2 | df | Prob>chi2 |
| --- | --- | --- | --- | --- | --- |
| Suspected Omicron |  | 0.1213 | 1.97 | 1 | 0.1607 |
| Male sex |  | -0.01886 | 0.05 | 1 | 0.8285 |
| Age category | 30-39 years | -0.06309 | 0.49 | 1 | 0.4818 |
|  | 40-49 years | 0.11636 | 1.72 | 1 | 0.1895 |
|  | 50-59 years | -0.04749 | 0.27 | 1 | 0.6053 |
|  | 60-69 years | -0.01227 | 0.02 | 1 | 0.8864 |
|  | ≥70 years | 0.06897 | 0.65 | 1 | 0.4216 |
| Comorbidities | HIV positive | -0.0443 | 0.26 | 1 | 0.608 |
|  | Diabetes | -0.03791 | 0.23 | 1 | 0.6311 |
|  | TB (current) | 0.0296 | 0.13 | 1 | 0.722 |
|  | HPT | -0.05891 | 0.49 | 1 | 0.4824 |
|  | CKD | 0.1688 | 4.6 | 1 | 0.0319 |
|  | COPD | -0.07967 | 0.87 | 1 | 0.3522 |
| Reinfection |  | 0.08018 | 0 | 1 | 1 |
| Vaccination | Partial | -0.03976 | 0.19 | 1 | 0.6656 |
|  | Complete | 0.18966 | 4.41 | 1 | 0.0358 |
| Global test |  |  | 19.51 | 16 | 0.2433 |

**Supplementary Figure 1:**
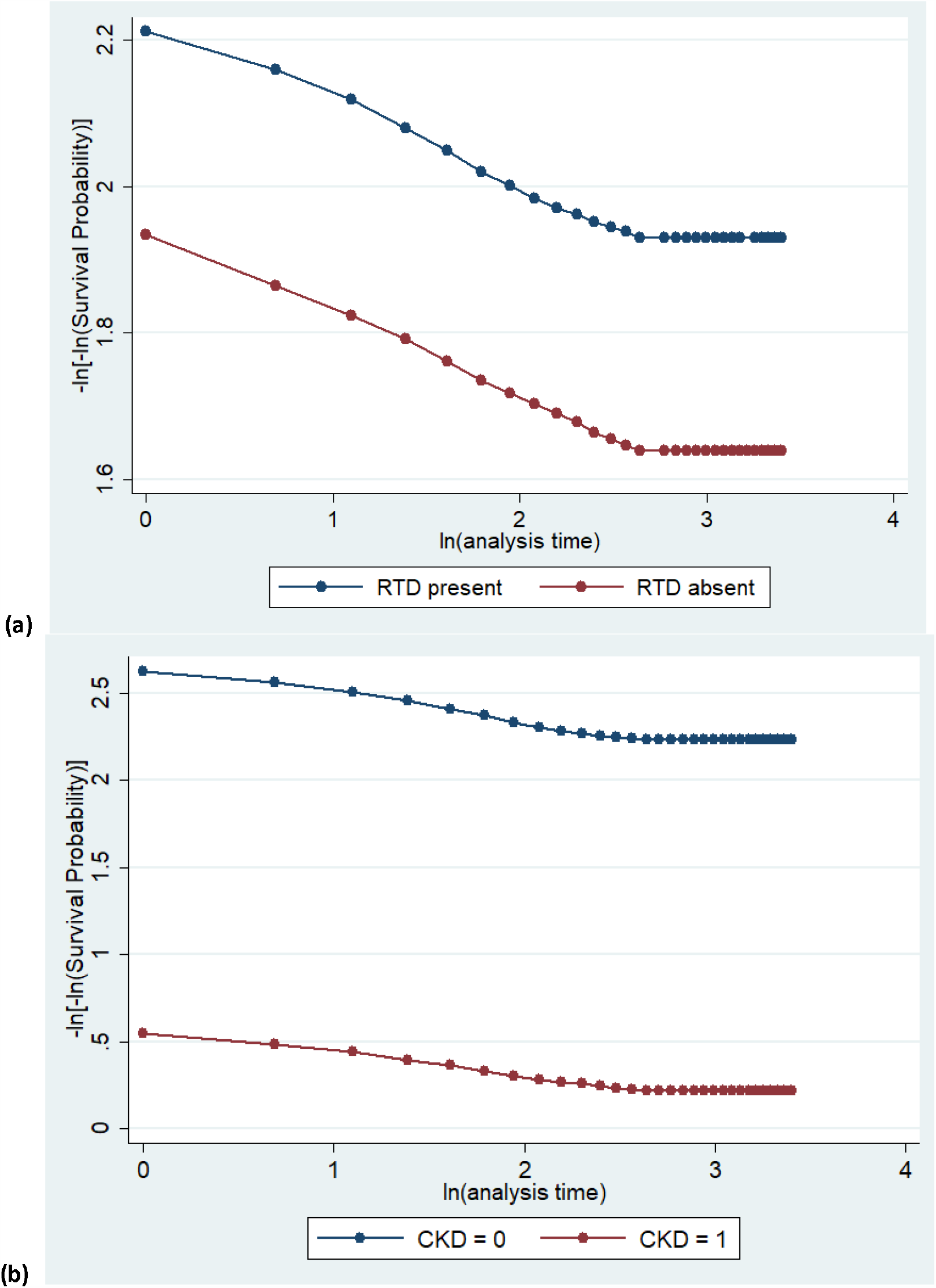
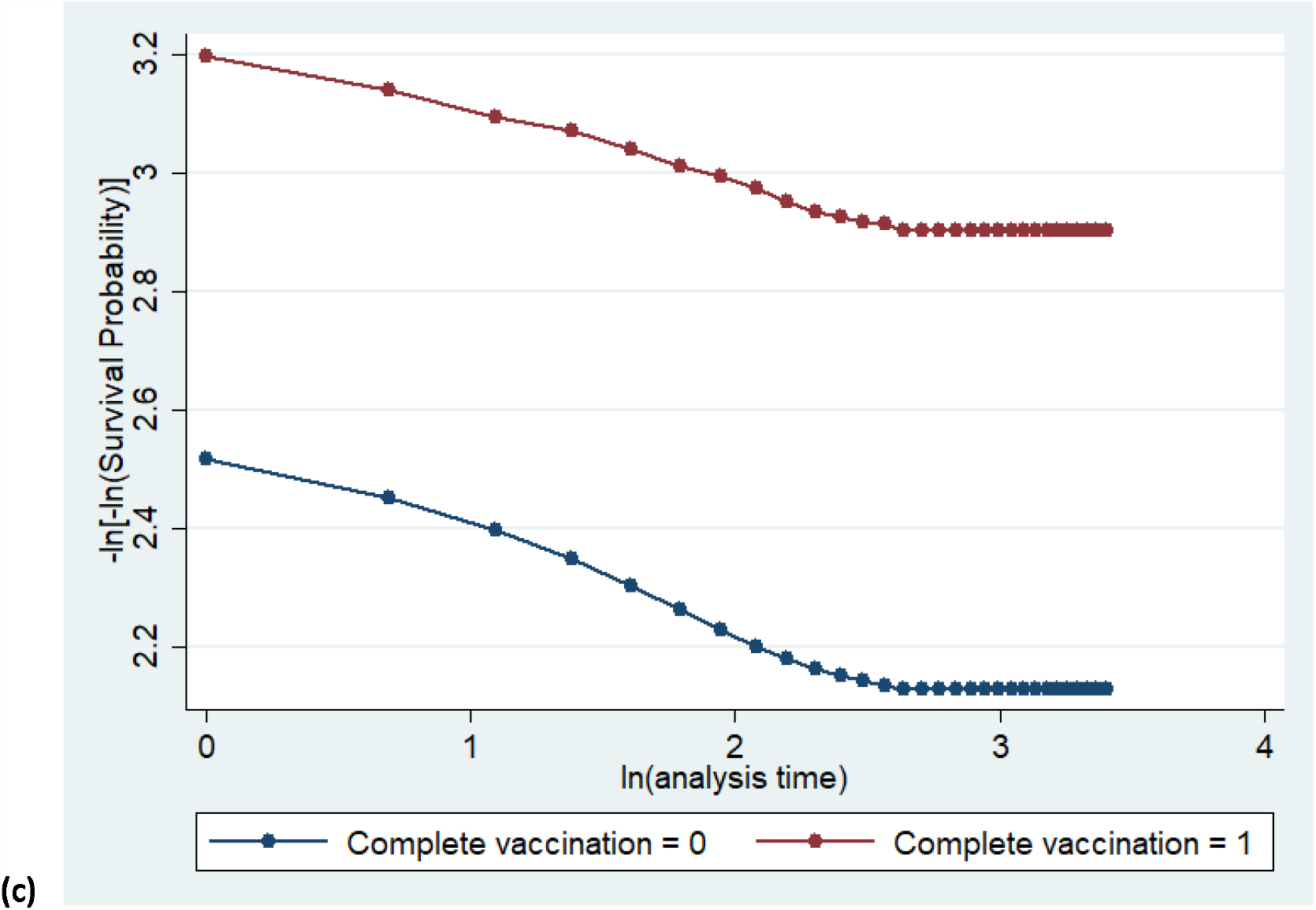
Log-log plots for the covariates of (a) RTD, (b) CKD and (c) complete vaccination.

