## Supplemental Figure 1 for "Assessing the clinical severity of the Omicron variant in the Western Cape Province, South Africa, using the diagnostic PCR proxy marker of RdRp target delay to distinguish between Omicron and Delta infections – a survival analysis"

(a)
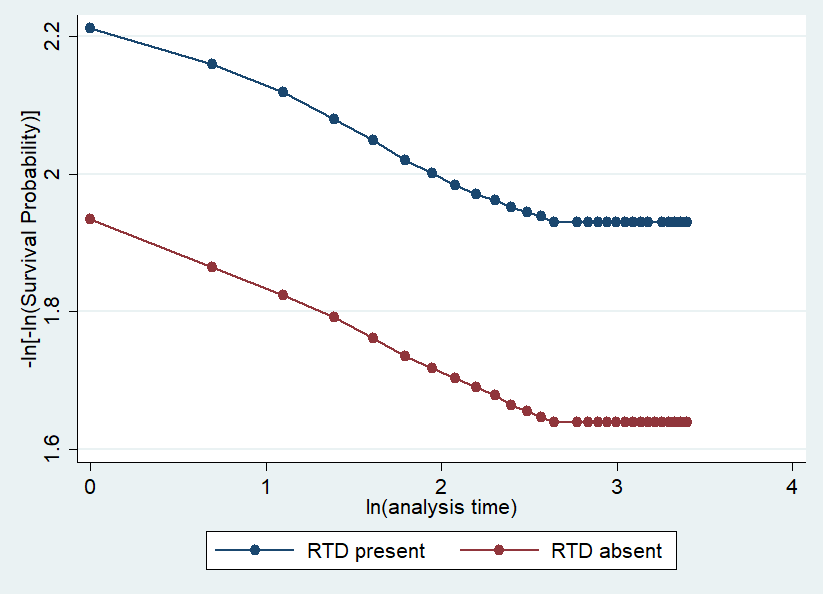

(b)
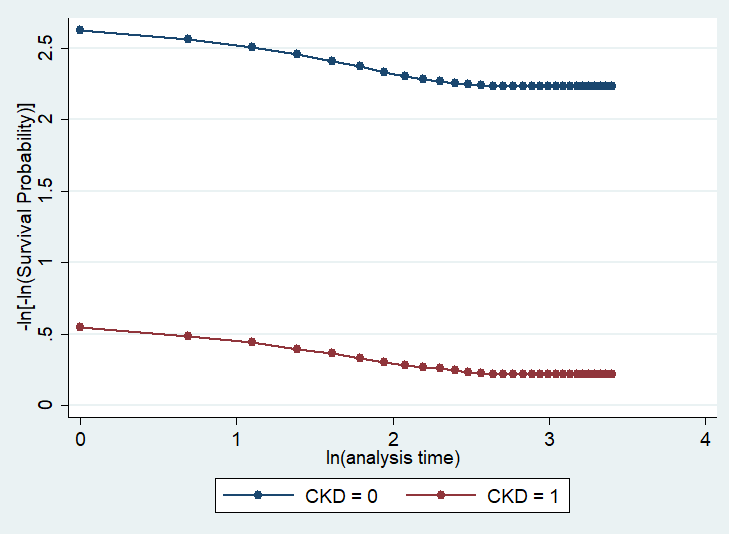

(c)
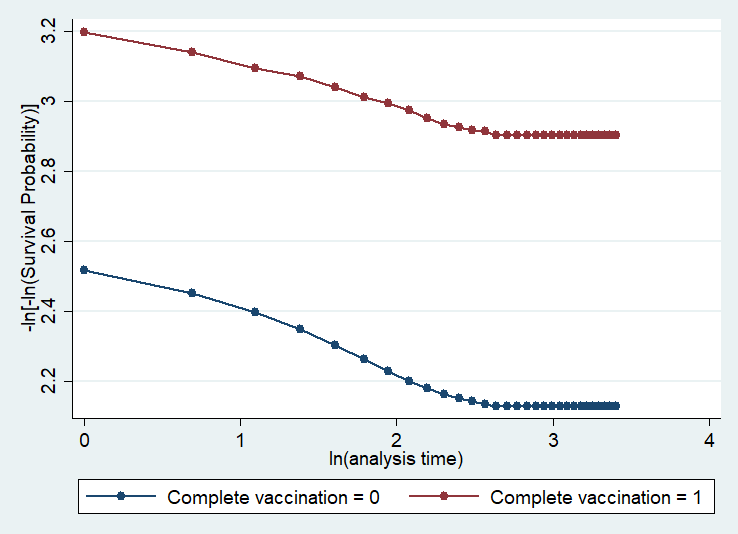

**Supplementary Figure 1: Log-log plots for the covariates of (a) RTD, (b) CKD and (c) complete vaccination.**
